# Saccharomyces boulardii as Adjunctive Therapy in Helicobacter pylori Eradication: A Randomized, Placebo-Controlled Trial

**DOI:** 10.64898/2026.11.09.26367596

**Authors:** Mohammad Jafari, Nahid Moradi, Hadi Abdolahi, Fatemeh Keshaei

## Abstract

**Background:** Helicobacter pylori (H. pylori) infection remains highly prevalent in Iran, where high antibiotic resistance can limit eradication efficacy. Saccharomyces boulardii (S. boulardii) may improve eradication and treatment tolerability, although evidence remains inconsistent. This trial evaluated adjunctive S. boulardii in a high-prevalence, high-resistance setting.

**Methods:** In this randomized, single-blind, placebo-controlled trial, 218 H. pylori–positive adults received 14 days of PPI, bismuth, amoxicillin, and clarithromycin (P-BAC) plus either S. boulardii (250 mg twice daily) or placebo. The primary outcome was H. pylori eradication assessed 30 days after treatment by stool antigen testing, analyzed by intention-to-treat (ITT) and per-protocol (PP) principles. Treatment-related adverse events were secondary outcomes. Trial registration: Iranian Registry of Clinical Trials, IRCT20230827059280N1.

**Results:** In the PP analysis, eradication was lower with S. boulardii than placebo (65.0% vs. 76.5%; RR = 0.849, 95% CI: 0.709–1.018), with a similar pattern in the ITT analysis (59.6% vs. 68.8%; RR = 0.867, 95% CI: 0.710–1.058). Neither analysis showed a statistically significant difference. Overall adverse events were significantly less frequent with S. boulardii (44.0% vs. 68.4%; RR = 0.644, 95% CI: 0.497–0.834; P < 0.001), with lower rates of bloating (4.0% vs. 23.5%; adjusted P < 0.001) and epigastric pain (4.0% vs. 14.3%; adjusted P = 0.041).

**Conclusion:** Adjunctive S. boulardii did not significantly improve H. pylori eradication with 14-day P-BAC therapy. However, it significantly reduced adverse events, particularly bloating and epigastric pain. Further multicenter trials incorporating individualized susceptibility testing are warranted.

## Introduction

Helicobacter pylori (H. pylori) remains a major global public health challenge, infecting an estimated 48.9% of the worldwide population.^1^ In Iran, this burden is even higher. The national prevalence is approximately 54% overall and 62% in the adult population, classifying the country as a high-prevalence region.^2^ Although the infection is usually asymptomatic, it can drive a wide range of gastroduodenal pathologies, including gastritis, peptic ulcer disease (PUD), gastric cancer, and gastric mucosa-associated lymphoid tissue (MALT) lymphoma.^3^ Moreover, H. pylori is classified as a group 1 carcinogen by the International Agency for Research on Cancer (IARC).^4^ 69.7% of gastric cancers worldwide, and 75.2% of cases in Iran are associated with its infection.^5^ Beyond the gastrointestinal tract, H. pylori also alters the gut microbiome and triggers systemic inflammation, contributing to various extra-gastric disorders.^6^ Therefore, successful H. pylori eradication is essential to prevent peptic ulcers, reduce cancer risks, and control these broader systemic conditions.

Choosing an optimal H. pylori eradication regimen depends on regional antibiotic resistance patterns and drug availability.^7^ Based on evidence from epidemiologically similar regions, our center utilizes a bismuth-based quadruple therapy consisting of a proton pump inhibitor (PPI), bismuth, amoxicillin, and clarithromycin (P-BAC) as the first-line protocol.^8,9^ However, antimicrobial resistance patterns suggest that the efficacy of this standard regimen will decline without therapeutic modifications. Local H. pylori resistance rates for clarithromycin and amoxicillin have reached 25.3% and 20.7%, respectively.^10^ Novel acid suppression strategies and alternative antibiotic regimens can improve eradication outcomes. However, their widespread implementation may be limited in some regions due to cost, availability, and concerns regarding antimicrobial resistance.^11,12^ Therefore, there is a need for accessible, cost-effective, and safe adjunctive strategies that could enhance the effectiveness of standard H. pylori eradication therapy.

Another major concern is the adverse events associated with H. pylori eradication regimens. In a large study of the European registry (Hp-EuReg), 37% of patients treated with the P-BAC regimen for 14 days experienced at least one adverse event. The frequent complications were metallic taste, diarrhea, nausea, vomiting, dyspepsia, heartburn, and abdominal pain.^13^ Other studies indicate that above 70% of patients receiving regimens containing amoxicillin and clarithromycin experience adverse events, most notably epigastric pain and taste disturbance.^14^ These antibiotics can also negatively impact the gut microbiome, leading to dysbiosis and increased susceptibility to opportunistic pathogens.^15^ Alterations in the gut microbiome impair intestinal permeability, modulate immune response, induce visceral hypersensitivity, and disrupt gut-brain signaling. All these mechanisms are associated with the development of dyspeptic symptoms, epigastric pain, and bloating.^16,17^ Using adjunctive agents may represent a strategy to reduce these adverse events and improve treatment tolerability.

Probiotics as an adjunctive therapy during H. pylori eradication may have several potential benefits, including direct pathogen suppression, preservation of the gut microbiome, and modulation of the immune response to mitigate inflammation.^18^ Saccharomyces boulardii (S. boulardii) is a non-pathogenic probiotic yeast. It has several potential mechanisms, such as inhibiting bacterial pathogens, reducing pro-inflammatory cytokines, and modulating the gut microbiome.^19^ S. boulardii’s fungal nature makes it resistant to antibiotics. So, unlike bacterial probiotics, it cannot contribute to the development of further resistance patterns. This feature is particularly important in settings with high antibiotic resistance.^20^ These combined properties could enhance H. pylori clearance, improve treatment tolerability, and reduce treatment-related adverse events.

While some previous studies suggest that S. boulardii may improve H. pylori eradication rates ^21^, others have failed to confirm this benefit.^22^ In contrast, there is stronger consensus regarding its positive role in reducing treatment-related adverse events. The specific adverse events affected vary significantly across trials, though.^21–23^ These controversies are likely due to variations among studied populations, particularly in regional antibiotic resistance patterns and H. pylori prevalence, as well as differences in underlying treatment protocols. To address these inconsistencies, we conducted a randomized, placebo-controlled clinical trial evaluating S. boulardii supplementation alongside a P-BAC eradication regimen in a region with high antibiotic resistance and H. pylori prevalence. This study aimed to determine the impact of S. boulardii on both eradication efficacy and adverse event rates under these specific regional conditions.

## Materials and Methods

### Study Design

This randomized, single-blind, parallel-group clinical trial was conducted at the Gastroenterology Clinic of Alzahra Hospital, Isfahan, Iran, between December 2023 and March 2025. This trial was originally designed as a three-arm study comparing placebo, S. boulardii, and rosuvastatin as adjuncts to standard H. pylori eradication therapy. The present article reports the pre-specified primary comparison between S. boulardii and placebo. We evaluated the second treatment arm (rosuvastatin) against the same placebo group and will report it in a separate publication. The ethics committee of Isfahan University of Medical Sciences approved the study protocol (IR.MUI.MED.REC.1402.164), and the trial was registered at the Iran Registry of Clinical Trials (IRCT20230827059280N1). The study was conducted in accordance with the principles of the Declaration of Helsinki.

### Participants

Patients referred to the Gastroenterology Clinic were screened for eligibility according to the following inclusion and exclusion criteria:

Adults aged 18 to 80 years presenting with an indication for H. pylori testing according to the 2017 ACG guidelines^24^, including PUD, MALT lymphoma, dyspepsia, planned long-term NSAID or aspirin therapy, history of early gastric cancer, or high risk of gastric cancer, underwent H. pylori stool antigen testing. Individuals with a positive stool antigen test were eligible for enrollment in the study.

Patients were excluded if they were pregnant or lactating, had known immunodeficiency, had used macrolides, probiotics, or statins within the preceding 3 months, had impaired liver function, defined as AST or ALT levels >5 times the upper limit of normal, or had a history of hypersensitivity to penicillin or any component of the study treatment regimen.

### Informed Consent and Withdrawals

All participants provided written informed consent. Participants were withdrawn from the study under the following situations: poor treatment compliance (e.g., failure to take the prescribed medication as instructed), failure to return for scheduled follow-up visits, voluntary withdrawal from the study, development of severe side effects such as anaphylaxis, or requirement for further treatment with probiotics, antibiotics, or PPIs after completion of the study regimen.

### Sample Size Calculation

Sample size was calculated separately for the planned pairwise comparisons of S. boulardii versus placebo and rosuvastatin versus placebo in the parent three-arm trial. Because both experimental comparisons shared a common placebo comparator, we estimated the expected eradication rate in the placebo group from a previous randomized trial in which patients received the same bismuth-containing quadruple therapy regimen (P-BAC) plus placebo.^25^

An absolute difference of 20% in eradication rate (85.45% vs. 65.45%) was considered the minimum clinically meaningful treatment effect, given the local healthcare setting, treatment costs, the high prevalence of antibiotic resistance, and the potential adverse events of adjunctive therapies. Using a two-sided α level of 0.05 and 90% power, the required sample size was calculated with the formula for comparing two independent proportions. The calculated sample size was subsequently increased by 10% to account for anticipated participant attrition, resulting in a target enrollment of 109 participants per group. This sample size was applied uniformly across all three study arms.

### Randomization and Blinding

Participants were allocated to the three study groups using a computer-generated block randomization sequence (block size of six, allocation ratio 1:1:1) generated using Random Allocation software by an investigator not involved in participant recruitment, treatment administration, or outcome assessment. The allocation codes (A, B, and C) were placed in the order generated into identical opaque, sealed, sequentially numbered envelopes. Code A represented the placebo group, code B the S. boulardii group, and code C the rosuvastatin group. The treating physician, who enrolled participants and dispensed medications, was not involved in sequence generation and was unaware of the block size. After enrollment, the corresponding envelope was opened, and the assigned intervention was provided.

The trial was single-blinded, with participant blinding. Study medications were prepared in visually identical capsules, and participants remained unaware of their allocation throughout the study. Laboratory personnel assessing the stool antigen test and the statistician were blinded to treatment allocation.

### Clinical Visits

Each participant was scheduled for the following visits and contacts:

1. Day 0 (baseline visit): Enrollment, eligibility confirmation, and collection of baseline characteristics, medical history, lifestyle factors, and anthropometric data retrieved from medical records using structured data collection forms.
2. Day 7 (telephone contact): Structured assessment of treatment-related adverse events, including epigastric pain, diarrhea, constipation, black stool, nausea, bloating, and taste disturbance.
3. Day 14 (end of treatment visit): Treatment adherence was verified at the day 14 visit through return of medication containers. Participants were classified as adherent only if empty containers for all prescribed medications were returned, confirming completion of the full course. Non-adherent participants were excluded from the per-protocol analysis. Treatment-related adverse events were reassessed using the same structured questionnaire.
4. Day 44 (final visit): Thirty days after completion of eradication therapy, H. pylori eradication was assessed using a monoclonal stool antigen test. Adverse events reported since the previous assessment were also recorded.

Participants were also permitted to return for evaluation in case of serious adverse events, such as hypersensitivity reactions, at any point during the study period.

### Treatment Protocol

All participants received the standard *H. pylori* P-BAC eradication regimen as follows: Esomeprazole 40 mg every 12 hours, clarithromycin 500 mg every 12 hours, amoxicillin 1000 mg every 12 hours, and bismuth subcitrate 120 mg every 6 hours for 14 days. Participants in the intervention group received capsules containing 250 mg of *S. boulardii* (equivalent to 5 billion viable cells; Sabular®, Farabiotic, Iran) every 12 hours, in addition to the standard therapy, for 14 days. The control group received identical placebo capsules matched in appearance, taste, and odor to the probiotic capsules every 12 hours. Patients were instructed not to use PPIs, antibiotics, or probiotics between the end of therapy and the final stool antigen test.

### Study Outcome

The primary outcome was the H. pylori eradication rate, assessed 30 days after completion of eradication therapy using a monoclonal stool antigen test. Eradication was defined as a negative stool antigen test result. Secondary outcomes included the incidence of treatment-related adverse events.

## Statistical Analysis

Statistical analyses were performed using Python (version 3.13.9) with the pandas, NumPy, SciPy, and statsmodels libraries. The normality of continuous variables was assessed using the Shapiro–Wilk test. Because the continuous variables were not normally distributed, we summarized them as median (Q1–Q3) and compared them between groups using the Mann–Whitney U test. Effect size (r) was reported for comparisons of baseline continuous variables. Categorical variables were compared using the chi-square test or Fisher’s exact test, as appropriate.

The primary outcome was analyzed according to both the intention-to-treat (ITT) and per-protocol (PP) principles. For the ITT analysis, all randomized participants were included. Participants with missing H. pylori eradication outcome data or those excluded from the PP analysis were considered treatment failures. To account for multiple comparisons, P-values for adverse-event outcomes were adjusted using the Benjamini–Hochberg false discovery rate (FDR) procedure. Treatment effects for the primary and secondary outcomes were expressed as relative risks (RRs) with 95% confidence intervals (CIs).

Treatment effects for the primary and secondary outcomes were expressed as relative risks (RRs) with 95% confidence intervals (CIs). Multivariable logistic regression was performed to estimate adjusted odds ratios (ORs) for treatment success and complications. Both full and reduced models were fitted, with all covariates in each model entered simultaneously. The full model included treatment group, age, sex, BMI, smoking, coffee consumption, diabetes, gastrointestinal disease, hypertension, and thyroid disease. The reduced model included treatment group, age, sex, and BMI to limit potential overfitting. As this manuscript reports the pre-specified comparison between the S. boulardii and placebo groups from the parent three-arm trial, all analyses were restricted to these two study arms. All statistical tests were two-sided, and a P-value <0.05 was considered statistically significant.

## Results

Among the participants enrolled in the parent three-arm trial, 218 were randomized to the placebo group (n = 109) or the S. boulardii group (n = 109), which constitute the treatment arms analyzed in the present study. During follow-up, 11 participants in the placebo group and 9 in the S. boulardii group were excluded from the per-protocol analysis primarily due to non-adherence to the prescribed regimen, loss to follow-up, or failure to provide a post-treatment stool antigen test. As a result, 198 participants (98 and 100, respectively) were included in the per-protocol analysis, whereas all 218 randomized participants were included in the intention-to-treat (ITT) analysis. Details of participant flow and reasons for study discontinuation are presented in Figure 1.

**Figure 1.**
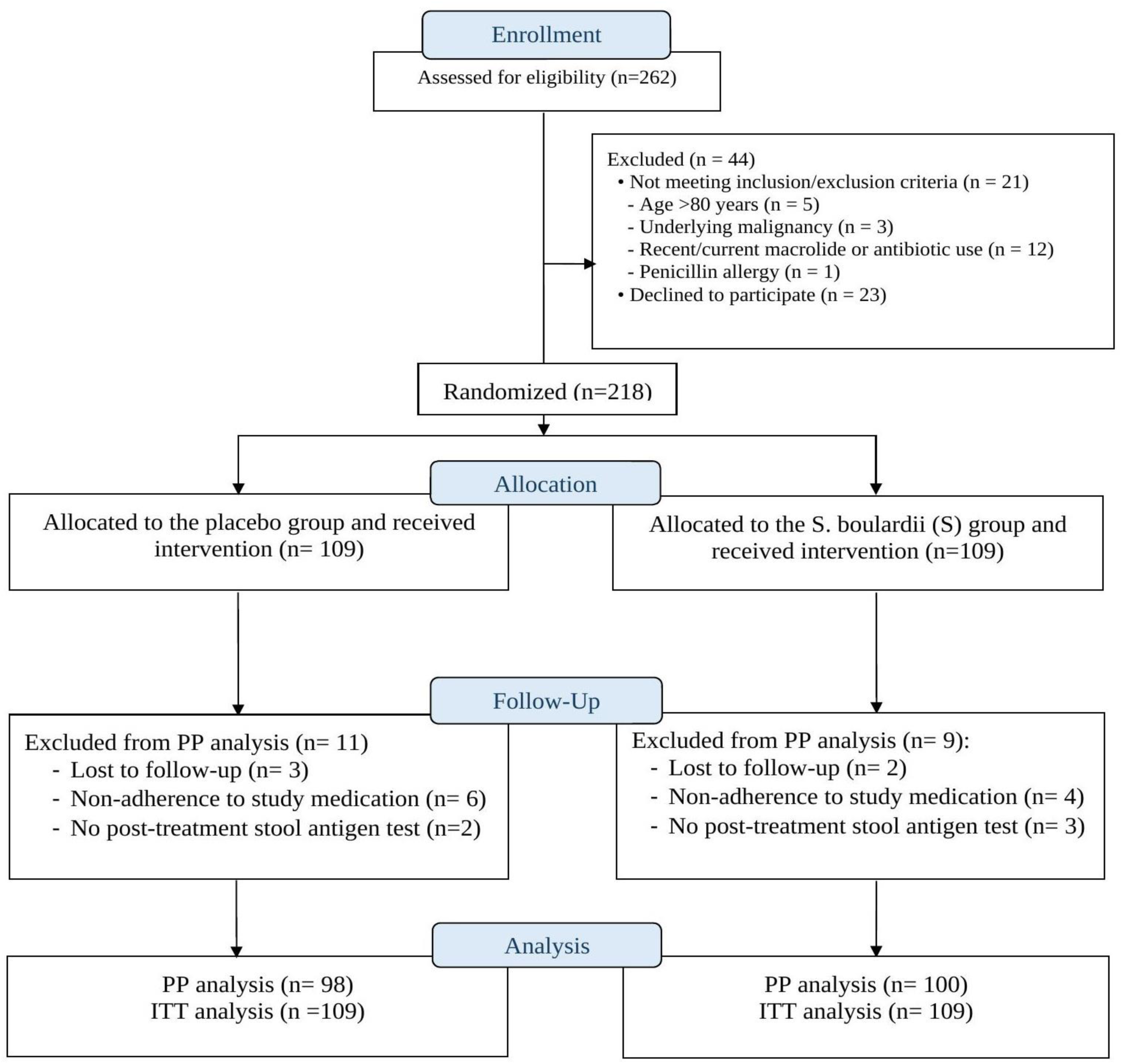
CONSORT flow diagram of participant enrollment, randomization, follow-up, and analysis in the placebo and S. boulardii groups. PP, per protocol; ITT, intention to treat.

Baseline demographic and clinical characteristics of participants, including age, sex, BMI, primary signs and symptoms, lifestyle factors, and underlying health conditions, are summarized in Table 1. The two groups had no statistically significant differences for any variable (P = [0.076–0.953], Table 1).

**Table 1.** Baseline demographic, lifestyle, and clinical characteristics of the placebo and S. boulardii groups.

| **Variable** | **Placebo (n = 98)** | **S. boulardii (n = 100)** | **Test** | **P-value** | **r** |
|---|---|---|---|---|---|
| **Baseline demographics** |  |  |  |  |  |
| Age, years | 47.00 (35.25–59.00) | 48.00 (39.50–58.00) | Mann-Whitney U | 0.606 | 0.037 |
| Height, cm | 167.50 (160.00–172.00) | 165.00 (160.00–173.00) | Mann-Whitney U | 0.953 | 0.004 |
| Weight, kg | 70.00 (63.00–79.50) | 70.00 (64.00–80.00) | Mann-Whitney U | 0.790 | 0.019 |
| BMI, kg/m<sup>2</sup> | 25.05 (23.53–27.30) | 25.57 (22.93–27.81) | Mann-Whitney U | 0.713 | 0.026 |
| Male | 37 (37.8%) | 47 (47.0%) | Chi-square | 0.188 | — |
| Female | 61 (62.2%) | 53 (53.0%) |  |  | — |
| **Lifestyle Factors** |  |  |  |  |  |
| Coffee consumption | 14 (14.3%) | 23 (23.0%) | Chi-square | 0.116 | — |
| Smoking | 13 (13.3%) | 23 (23.0%) | Chi-square | 0.076 | — |
| Alcohol consumption | 2 (2.0%) | 6 (6.0%) | Fisher | 0.279 | — |
| **Primary signs and symptoms** |  |  |  |  |  |
| Dyspepsia | 57 (58.2%) | 53 (53.0%) | Chi-square | 0.465 | — |
| Heartburn | 29 (29.6%) | 23 (23.0%) | Chi-square | 0.292 | — |
| Gastrointestinal bleeding | 13 (13.3%) | 20 (20.0%) | Chi-square | 0.204 | — |
| Diarrhea | 9 (9.2%) | 8 (8.0%) | Chi-square | 0.766 | — |
| Other | 2 (2.0%) | 0 (0.0%) | Fisher | 0.244 | — |
| **Underlying health conditions** |  |  |  |  |  |
| Hypertension | 23 (23.5%) | 23 (23.0%) | Chi-square | 0.938 | — |
| Diabetes | 16 (16.3%) | 18 (18.0%) | Chi-square | 0.755 | — |
| Thyroid disease | 11 (11.2%) | 12 (12.0%) | Chi-square | 0.865 | — |
| Gastrointestinal disease | 9 (9.2%) | 6 (6.0%) | Chi-square | 0.397 | — |
| Cardiovascular disease | 0 (0.0%) | 4 (4.0%) | Fisher | 0.121 | — |

*BMI, body mass index; IQR, interquartile range; r, effect size.*

The H. pylori eradication rate was numerically lower in the S. boulardii group than in the placebo group in both the per-protocol analysis (65.0% vs. 76.5%; RR = 0.849, 95% CI: 0.709–1.018) and the intention-to-treat analysis (59.6% vs. 68.8%; RR = 0.867, 95% CI: 0.710–1.058). However, the difference was not statistically significant in either analysis (P = 0.075, P = 0.158, respectively, Table 2, Figure 2)

**Figure 2.**
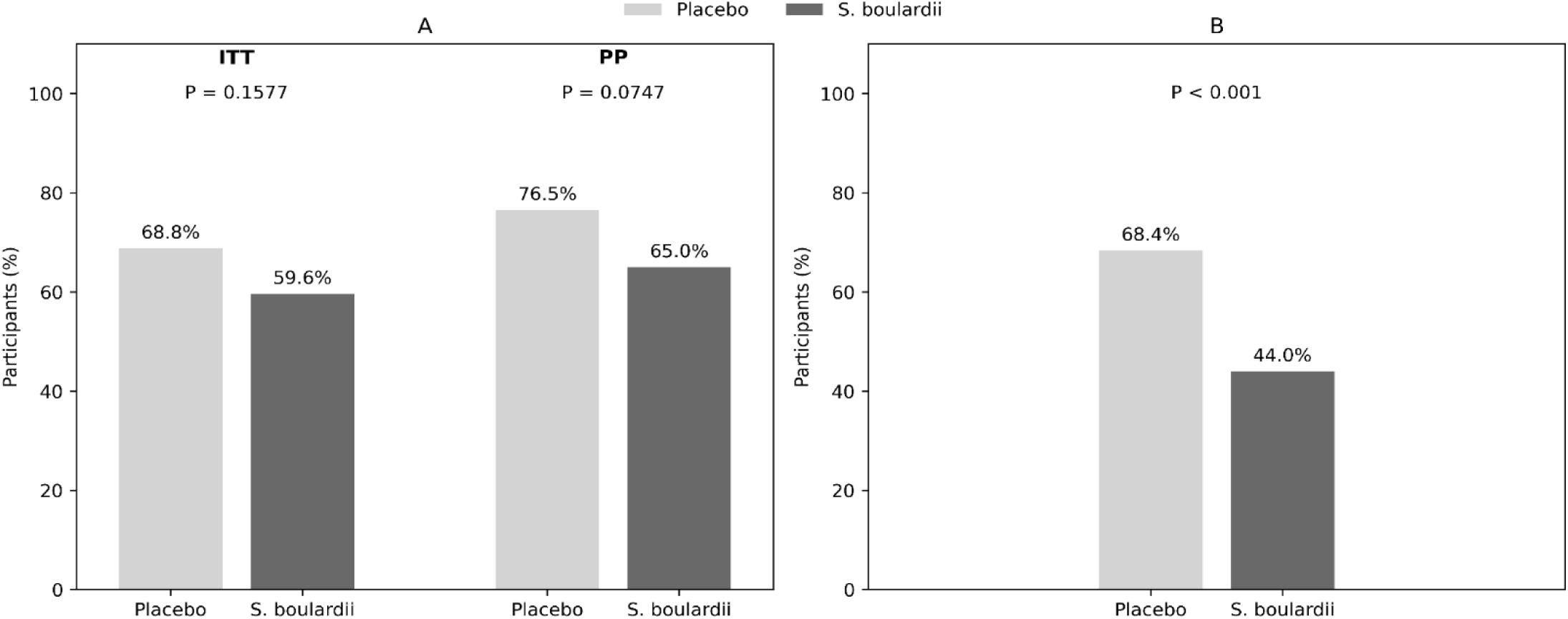
Comparison of Helicobacter pylori eradication rates and treatment-related adverse events between the Saccharomyces boulardii and control groups. (A) Eradication rates in the intention-to-treat (ITT) and per-protocol (PP) populations. (B) Proportion of partic experiencing treatment-related adverse events in each group. P-values are shown above the bars.

**Table 2.** Eradication rates in the Placebo and S. boulardii groups (ITT and PP analysis)

| **Outcome** | **Analysis** | **Probiotic n (%)** | **Control n (%)** | **RR** | **95% CI** | **P-value** |
|---|---|---|---|---|---|---|
| Successful eradication | Per-protocol | 65/100 (65.0%) | 75/98 (76.5%) | 0.849 | 0.709 - 1.018 | 0.075 |
|  | ITT | 65/109 (59.6%) | 75/109 (68.8%) | 0.867 | 0.710 - 1.058 | 0.158 |

*ITT, intention to treat; PP, per protocol; RR, relative risk; CI, confidence interval.*

The overall incidence of adverse events was significantly lower in the S. boulardii group than in the placebo group (44.0% vs. 68.4%; RR = 0.644; 95% CI: 0.497–0.834; P < 0.001). The absolute risk reduction was 24.4% with a number needed to treat (NNT) of 5. Among the seven individual adverse events assessed, bloating (4.0% vs. 23.5%; adjusted P < 0.001) and epigastric pain (4.0% vs. 14.3%; adjusted P = 0.041) remained significantly less frequent in the S. boulardii group after Benjamini–Hochberg correction for multiple comparisons. No significant differences were observed between groups for the remaining adverse events, including bitter taste, black stool, nausea, diarrhea, and constipation, after adjustment (Table 3). Figure 3 illustrates the relative risks and 95% confidence intervals for overall and individual adverse events.

**Figure 3.**
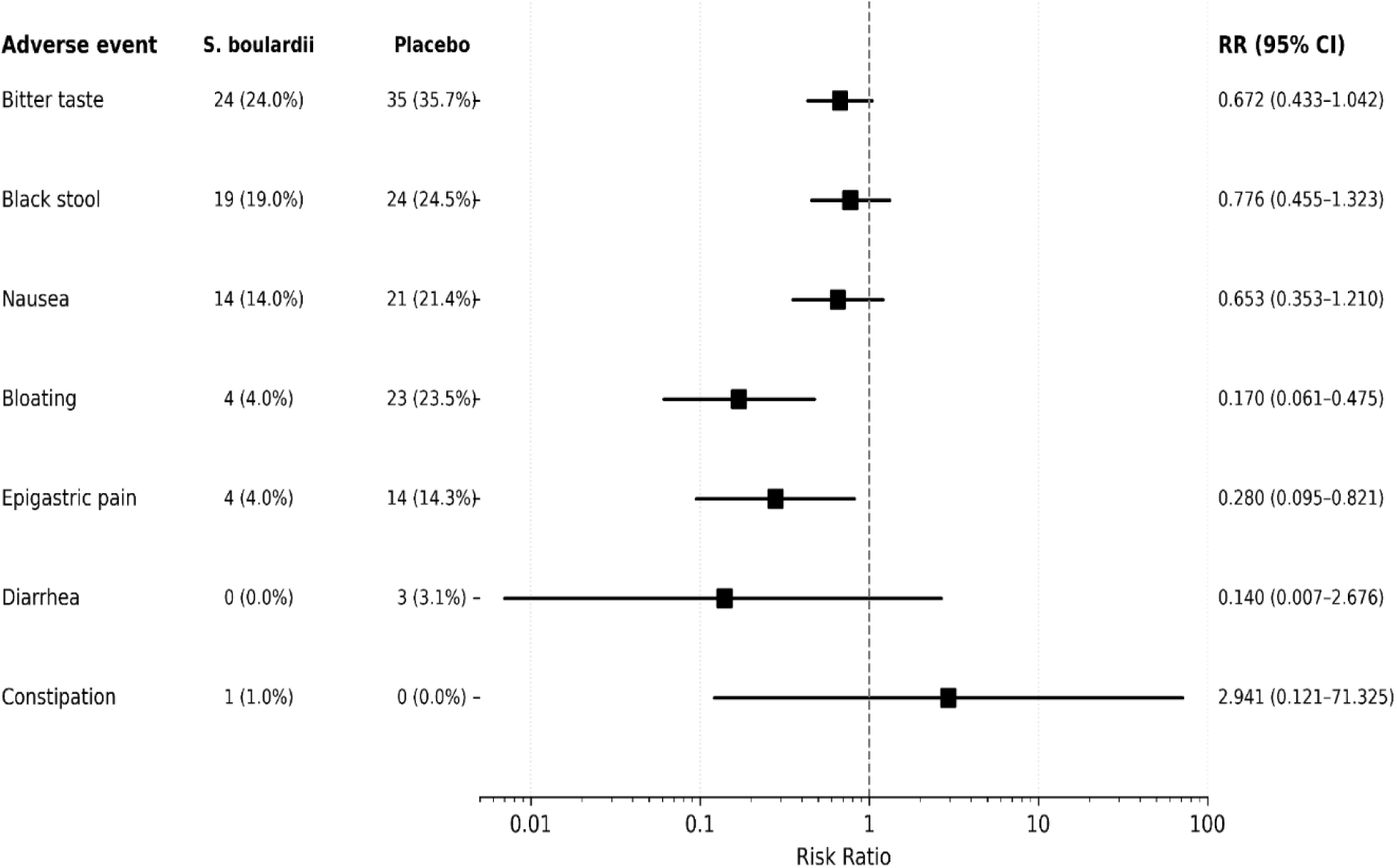
Forest plot of risk ratios for treatment-related adverse events in the placebo and S. boulardii groups. RR, relative risk; CI, confidence interval.

**Table 3.** Overall and individual treatment-related adverse events in the placebo and S. boulardii groups.

| **Adverse event** | **S. boulardii n (%)** | **Control n (%)** | **Test** | **RR** | **95% CI** | **Unadjusted P-value** | **FDR-adjusted P-value** |
|---|---|---|---|---|---|---|---|
| Bitter taste | 24 (24.0%) | 35 (35.7%) | Chi-square | 0.672 | 0.433 - 1.042 | 0.072 | 0.167 |
| Black stool | 19 (19.0%) | 24 (24.5%) | Chi-square | 0.776 | 0.455 - 1.323 | 0.345 | 0.407 |
| Nausea | 14 (14.0%) | 21 (21.4%) | Chi-square | 0.653 | 0.353 - 1.210 | 0.171 | 0.239 |
| Bloating | 4 (4.0%) | 23 (23.5%) | Chi-square | 0.17 | 0.061 - 0.475 | <0.001 | <0.001 |
| Epigastric pain | 4 (4.0%) | 14 (14.3%) | Chi-square | 0.28 | 0.095 - 0.821 | 0.012 | 0.041 |
| Diarrhea | 0 (0.0%) | 3 (3.1%) | Fisher | 0.14 | 0.007 - 2.676 | 0.119 | 0.209 |
| Constipation | 1 (1.0%) | 0 (0.0%) | Fisher | 2.941 | 0.121 - 71.325 | 1 | 1 |
| Overall | 44 (44.0%) | 67 (68.4%) | Chi-square | 0.644 | 0.497 – 0.834 | <0.001 | — |

*RR, relative risk; CI, confidence interval; FDR, false discovery rate (P-values were adjusted using the Benjamini–Hochberg procedure).*

To account for potential confounders, multivariable logistic regression analysis was performed to evaluate the association between the treatment group and H. pylori eradication. Consistent with the unadjusted analysis, S. boulardii group was not significantly associated with eradication success after adjustment for age, sex, and BMI (adjusted OR = 0.555, 95% CI: 0.296–1.042, P = 0.067), nor after further adjustment for comorbidities (diabetes mellitus, gastrointestinal disease, hypertension, and thyroid disease) and lifestyle factors (smoking and coffee consumption) (adjusted OR = 0.551, 95% CI: 0.288–1.054, P = 0.072). Detailed results are provided in Supplementary Table S1.

Multivariable logistic regression also showed that S. boulardii treatment was associated with significantly lower odds of overall treatment-related adverse events. This association remained significant after adjustment for age, sex, and BMI (adjusted OR = 0.364, 95% CI: 0.203–0.653, P<0.001) and after further adjustment for comorbidities and lifestyle factors (adjusted OR = 0.324, 95% CI: 0.176–0.594, P<0.001). Detailed results are provided in Supplementary Tables S2.

## Discussion

This randomized, placebo-controlled trial evaluated the efficacy and safety of S. boulardii supplementation in a region with a high prevalence of H. pylori infection and substantial antimicrobial resistance. Adjunctive S. boulardii administered with a 14-day P-BAC regimen did not significantly improve H. pylori eradication. In fact, eradication rates were numerically lower in the S. boulardii group than in the placebo group in both per-protocol (65.0% vs. 76.5%) and intention-to-treat (59.6% vs. 68.8%) analyses. However, these differences were not statistically significant (P= 0.075,0.158, respectively). The eradication rates in both groups remained below the 90% threshold recommended by the Maastricht VI/Florence Consensus for empirical therapy. ^26^ This finding may indirectly reflect a high prevalence of antimicrobial resistance previously reported in our region.^10^ Although the difference in eradication rates was not statistically significant, the study may have limited power to detect effects smaller than the prespecified 20-percentage-point difference. Therefore, a smaller treatment effect cannot be excluded. In contrast, S. boulardii supplementation significantly reduced treatment-related adverse events (44.0% vs. 68.4%, P<0.001). This improvement remained significant after adjustment for demographic characteristics and lifestyle factors. Lower rates of bloating and epigastric pain primarily drove the reduction in adverse events.

### Eradication rate

Several meta-analyses published in 2025 reported a beneficial effect of adjunctive S. boulardii on H. pylori eradication.^23,27,28^ However, given the limited number of relevant randomized controlled trials, these reviews include overlapping primary studies and should not necessarily be interpreted as fully independent confirmations of the same effect. Another meta-analysis published in 2024, restricted to trials evaluating S. boulardii in combination with bismuth-based quadruple therapies, similarly found a statistically significant, albeit modest, improvement in the intention-to-treat analysis (RR 1.05, 95% CI 1.00–1.10, P = 0.03), whereas the per-protocol analysis did not reach statistical significance (RR 1.03, 95% CI 1.00–1.06, P = 0.07).^22^

In contrast, many individual RCTs examining adjunctive S. boulardii have not demonstrated a statistically significant effect on eradication when analyzed separately.^29–32^ Notably, real-world data from the Hp-EuReg registry, specifically evaluating the P-BAC regimen, reported a significantly lower eradication rate with adjunctive S. boulardii than with standard therapy alone (91.7% vs. 94.2%, P = 0.028).^33^ Although observational data cannot establish causality, the direction of this association is consistent with the numerically lower eradication rates observed in our trial. The pattern of significant pooled estimates alongside largely non-significant individual RCTs may reflect a combination of a modest treatment effect, limited statistical power in individual trials, heterogeneity across studies in patient populations, eradication regimens, S. boulardii dose and duration, and study settings. Our findings should therefore be interpreted in this context. We observed numerically lower eradication rates with S. boulardii (RR 0.849, 95% CI 0.709–1.018 in the per-protocol analysis RR 0.867, 95% CI 0.710–1.058 in the intention-to-treat analysis). However, the confidence intervals crossed unity in both analyses, and these results do not provide evidence of a statistically significant detrimental effect. At the same time, the relatively wide confidence intervals indicate that a small beneficial or detrimental effect cannot be excluded. Another potential explanation relates to the timing of probiotic administration. A previous meta-analysis found that probiotics administered before or after eradication therapy were associated with improved eradication rates, whereas no significant benefit was observed when probiotics were administered concurrently with eradication therapy (RR 1.04, 95% CI 0.92–1.19).^34^ As S. boulardii was administered concurrently with P-BAC in our study, the timing of supplementation may have influenced its effect on eradication.

### Adverse events

Our finding regarding lower incidence of overall adverse events in the S. boulardii group (44.0% vs. 68.4%, P < 0.001) is consistent with previous clinical trials and registry data evaluating the effect of adjunctive S. boulardii with the P-BAC regimen. However, the specific adverse events affected have varied across studies. Naghibzadeh et al. reported reductions in nausea, diarrhea, headache, abdominal pain, and anxiety.^35^ Zhao et al. reported a reduction in abdominal distention and diarrhea.^29^ The HP-EuReg registry reported lower rates of dysgeusia, diarrhea, nausea, dyspepsia, heartburn, asthenia, and anorexia (all P < 0.05).^33^ Our reduction in epigastric pain (4% vs. 14.3%, adjusted P = 0.041) is in line with the lower rates of abdominal pain, dyspepsia, and heartburn reported in these studies, although these outcomes are not directly equivalent. Similarly, our reduction in bloating (4.0% vs. 23.5%, adjusted P < 0.001) is consistent with meta-analyses reporting decreased abdominal distention (pooled RR = 0.39, 95% CI: 0.26–0.59; pooled RR = 0.41, 95% CI: 0.23–0.72)^22,23^, although bloating and abdominal distension are not directly equivalent. In contrast, unlike previous reports, we found no significant differences in nausea, constipation, diarrhea, or taste disturbance (all P > 0.05).^22,23,33,35^

A previous meta-analysis found differences in the effects of probiotics across populations, probiotic preparations, and treatment durations for adverse events, particularly diarrhea and abdominal bloating.^36^ Moreover, another meta-analysis of H. pylori eradication regimens reported substantial heterogeneity in overall adverse-event incidence across studies, which was attributed to differences in populations, treatment regimens, and adverse-event reporting standards.^37^ These factors may explain the heterogeneity in different studies, including ours.

### Underlying mechanisms

A possible explanation for our finding is the effect of S. boulardii on the interaction between H. pylori and host cells. In an in vitro study, S. boulardii prevented H. pylori adhesion to duodenal cells, whereas this effect was not observed in gastric cells.^38^ Other mechanisms, such as competition for nutrients and immune modulation, may also reduce the bacterial burden rather than a direct bactericidal effect.^39^ This may explain why these effects did not translate into improved H. pylori eradication in our study. S. boulardii may reduce adverse events through several mechanisms. It can strengthen intestinal epithelial junctions, modulate the immune response, and help maintain the gut microbiome during antibiotic therapy. ^40,41^ Clinical studies have also shown that S. boulardii may alleviate symptoms such as bloating and abdominal pain in patients with antibiotic-associated alterations in the gut microbiome.^42^ Taken together, these mechanisms may explain why S. boulardii improved treatment tolerability without significantly affecting H. pylori eradication.

### Clinical relevance

These findings may be particularly relevant in high-resistance, high-prevalence settings such as Iran. As discussed earlier, recent data indicate clarithromycin and amoxicillin resistance rates of 25.3% and 20.7%, respectively.^10^ These resistance rates may limit the efficacy of the P-BAC regimen. In this context, the lack of a significant improvement in eradication with S. boulardii should be considered alongside its significant reduction in treatment-related adverse events. Reduced adverse effects do not necessarily translate into improved adherence. However, better treatment tolerability may still represent a clinically relevant benefit. This may be particularly important for symptoms such as bloating and epigastric pain, which were less frequently reported in the probiotic group in our study. So, even without a demonstrated eradication benefit, S. boulardii may have a potential role in improving the tolerability of eradication therapy in high-resistance settings. Whether this reduction in treatment-related adverse effects improves patient-reported treatment experience or other meaningful clinical outcomes in routine practice remains to be determined. Future studies should evaluate the effects of S. boulardii alongside different H. pylori eradication regimens and acid-suppressive strategies. In particular, studies using P-CAB-based regimens instead of PPI-based regimens may help determine whether the observed effect on treatment tolerability is consistent across different eradication strategies.

### Strengths and limitations

This trial has several strengths. These include its randomized, placebo-controlled design and a prospectively determined sample size based on a prespecified 20-percentage-point difference. Both intention-to-treat and per-protocol analyses were performed. We assessed adverse events at multiple time points during and after treatment, including days 7, 14, and 44. The statistical analysis also included Benjamini–Hochberg correction for multiple comparisons and multivariable adjustment for relevant confounders. In addition, the study provides clinical data from a high-prevalence, high-resistance setting, which may add to the available evidence on the use of S. boulardii in different regional contexts.

Several limitations should also be acknowledged. First, the trial was single-blind, with participants blinded to treatment allocation, whereas the treating physician was not. This may have introduced performance or assessment bias. However, laboratory personnel and the statistician remained blinded to treatment allocation. Second, the study did not perform individual antibiotic susceptibility testing. Therefore, the resistance profile of individual participants could not be determined, and the findings were interpreted in the context of available regional resistance data. Third, the study was conducted at a single center, which may limit the generalizability of the findings to other populations and clinical settings. Fourth, the sample size was calculated to detect a prespecified 20-percentage-point difference in eradication rates. Therefore, the study may have had limited power to detect smaller differences.

## Conclusion

In this setting with high resistance and high prevalence, adjunctive S. boulardii did not significantly improve H. pylori eradication when added to 14-day P-BAC therapy. Our study was designed to detect a prespecified 20-percentage-point difference but may have had limited power to detect smaller effects. S. boulardii significantly reduced treatment-related adverse events, particularly bloating and epigastric pain. Therefore, its main clinical benefit may be related to treatment tolerability rather than eradication efficacy. Further multicenter trials with individualized susceptibility testing, other main eradication regimens, and alternative dosing schedules are warranted.

## Data Availability

The original data contain individual-level participant information and are not publicly available due to ethical and privacy considerations. De-identified data may be made available by the corresponding author upon reasonable request, subject to applicable ethical and institutional requirements. The statistical analysis code is publicly available on GitHub at: https://github.com/nahid-moradi/H.pylori_S.boulardii_Analysis.

## Acknowledgements

This study was funded by Isfahan University of Medical Sciences.

The authors used ChatGPT (OpenAI, 2026) and Claude (Anthropic, 2026) to assist with manuscript preparation, including language editing, proofreading, refinement of statistical wording, and assistance with the presentation and explanation of technical and statistical content. The AI tools were also used to assist with debugging statistical analysis code. The authors executed and verified all statistical analyses. All AI-assisted suggestions and outputs were critically reviewed by the authors. The authors made all final decisions regarding the manuscript, analyses, interpretation, and conclusions and take full responsibility for the content of the final work.

---

**Supplementary Table S1.** Logistic regression analysis of factors associated with H. pylori eradication.

| **Variable** | **Full model** |  |  | **Reduced model** |  |  |
|---|---|---|---|---|---|---|
|  | **Adjusted OR** | **95% CI** | **P-value** | **Adjusted OR** | **95% CI** | **P-value** |
| Group (placebo/ S. boulardii) | 0.551 | 0.288 - 1.054 | 0.072 | 0.555 | 0.296 – 1.042 | 0.067 |
| Age | 1.018 | 0.989 - 1.048 | 0.225 | 1.018 | 0.994 – 1.043 | 0.140 |
| BMI | 0.985 | 0.901 - 1.076 | 0.735 | 0.989 | 0.907 – 1.079 | 0.805 |
| Gender | 0.992 | 0.482 - 2.041 | 0.982 | 0.983 | 0.523 – 1.845 | 0.957 |
| Smoking | 1.493 | 0.545 - 4.086 | 0.435 | - | - | - |
| Coffee consumption | 0.654 | 0.258 - 1.655 | 0.370 | - | - | - |
| Diabetes | 1.265 | 0.463 - 3.454 | 0.647 | - | - | - |
| Gastrointestinal disease | 1.878 | 0.440 - 8.009 | 0.394 | - | - | - |
| Hypertension | 0.821 | 0.327 - 2.063 | 0.675 | - | - | - |
| Thyroid disease | 2.506 | 0.765 - 8.207 | 0.129 | - | - | - |

*OR, odds ratio; CI, confidence interval; BMI, body mass index*

**Supplementary Table S2.** Logistic regression analysis of factors associated with treatment-related adverse effects.

| **Variable** | **Full model** |  |  | **Reduced model** |  |  |
|---|---|---|---|---|---|---|
|  | **Adjusted OR** | **95% CI** | **P-value** | **Adjusted OR** | **95% CI** | **P-value** |
| Group (placebo/ S. boulardii) | 0.324 | 0.176 - 0.594 | <0.001 | 0.364 | 0.203 – 0.653 | <0.001 |
| Age | 1.005 | 0.978 - 1.032 | 0.725 | 1.003 | 0.981 – 1.025 | 0.788 |
| BMI | 1.02 | 0.938 - 1.108 | 0.646 | 1.018 | 0.938 – 1.104 | 0.675 |
| Gender | 1.441 | 0.727 - 2.855 | 0.296 | 1.091 | 0.606 – 1.963 | 0.772 |
| Smoking | 1.907 | 0.750 - 4.850 | 0.175 | - | - | - |
| Coffee consumption | 1.546 | 0.626 - 3.823 | 0.345 | - | - | - |
| Diabetes | 0.979 | 0.398 - 2.407 | 0.963 | - | - | - |
| Gastrointestinal disease | 0.518 | 0.153 - 1.753 | 0.291 | - | - | - |
| Hypertension | 1.164 | 0.496 - 2.729 | 0.727 | - | - | - |
| Thyroid disease | 1.019 | 0.391 - 2.654 | 0.970 | - | - | - |

*OR, odds ratio; CI, confidence interval; BMI, body mass index*

